# Genetic liability to schizophrenia is associated with exposure to traumatic events in childhood

**DOI:** 10.1101/19006577

**Authors:** Hannah M Sallis, Jazz Croft, Alexandra Havdahl, Hannah J Jones, Erin C Dunn, George Davey Smith, Stanley Zammit, Marcus R Munafò

## Abstract

**Background:** There is a wealth of literature on the observed association between childhood trauma and psychotic illness. However, the relationship between childhood trauma and psychosis is complex and could be explained, in part, by gene-environment correlation.

**Methods:** The association between schizophrenia polygenic scores (PGS) and experiencing childhood trauma was investigated using data from the Avon Longitudinal Study of Parents and Children (ALSPAC) and the Norwegian Mother, Father and Child Cohort Study (MoBa). Schizophrenia PGS were derived in each cohort for children, mothers, and fathers where genetic data were available. Measures of trauma exposure were derived based on data collected throughout childhood and adolescence (0-17 years; ALSPAC) and at age 8 years (MoBa).

**Results:** Within ALSPAC, we found a positive association between schizophrenia PGS and exposure to trauma across childhood and adolescence; effect sizes were consistent for both child or maternal PGS. We found evidence of an association between the schizophrenia PGS and the majority of trauma subtypes investigated, with the exception of bullying. These results were comparable in MoBa. Within ALSPAC, genetic liability to a range of additional psychiatric traits was also associated with a greater trauma exposure.

**Conclusions:** Results from two international birth cohorts indicate that genetic liability for a range of psychiatric traits is associated with experiencing childhood trauma. GWAS of psychiatric phenotypes may also reflect risk factors for these phenotypes. Our findings also suggest that youth at higher genetic risk might require greater resources/support to ensure they grow-up in a healthy environment.

## Introduction

There is a wealth of literature from observational studies showing an association between childhood trauma and psychotic illness. Recent studies have suggested that exposure to interpersonal violence or neglect in childhood can increase the risk of psychotic symptoms by 2-3 times (van Dam *et al*., 2012; Varese *et al*., 2012; Trotta, Murray and Fisher, 2015; Cunningham, Hoy and Shannon, 2016; McGrath *et al*., 2017). These findings suggest that childhood trauma is a causal environmental risk factor for both sub-clinical psychotic symptoms and psychotic disorder. However, the relationship between childhood trauma and psychosis is complex and could be explained, in part, by gene-environment correlation.

Gene-environment correlation reflects the association between an individual’s genotype and their environment (Jaffee and Price, 2008). There are three types of gene-environment correlation, commonly referred to as: passive, evocative (or reactive), and active (or selective) (Jaffee and Price, 2008). In each case, there is an observable association between genotype and environment, although the presumed mechanisms underlying these correlations are distinct. *Passive gene-environment correlation* occurs when there is an association between genetic information passed from parent to child, and the environment in which the child is raised. For example, it could be that parents with a genetic predisposition to psychosis may be more liable to behaviours that create an environment in which the child is subject to exposure to traumatic stress. In this case, as for evocative and active gene-environment correlation, trauma would be a marker for genetic risk of psychosis, not necessarily a cause of psychosis (Jaffee and Price, 2008). In addition to passive gene-environment correlation, parental genotype can also influence child phenotype via the genetic information that is *not* transmitted. This process is known as dynastic effects, where parent phenotype can influence their offspring’s outcomes (Kong *et al*., 2018; Brumpton *et al*., 2019). *Evocative gene-environment correlation* is the association between an individual’s genetic predisposition to a certain behaviour, and reactions of others to that behaviour. For example, the child may exhibit traits that elicit reactions of others around them – e.g. harsher parenting or victimisation by peers. *Active gene-environment correlation* occurs when an individual seeks out a particular environment based on their genetic information. For example, individuals with increased genetic predisposition to sensation seeking behaviours may be more likely to seek out riskier environments, such as associating with individuals who misuse substances, and hence have increased risk of substance misuse themselves. In our psychosis example, children with certain traits may seek out environments in which they may be more vulnerable to exposure to traumatic events. The relative importance of each type of correlation is presumed to change with development, with passive most influential earlier in life and active becoming more important as individuals begin to select their own environment (Scarr and McCartney, 1983).

Building from these insights, we hypothesized that the observed association between childhood trauma and psychosis is due, in part, to a gene-environment correlation. In all three situations, an association between the occurrence of psychosis (as a result of increased genetic liability) and trauma could occur as a result of gene-environment correlation, meaning without trauma being causal. However, it is also possible that gene-environment correlation can be part of a causal chain, a fact utilised by a technique known as Mendelian randomization which relies on such correlations to infer causality (Davey Smith and Ebrahim, 2005; Gage *et al*., 2016). Whilst a number of lines of evidence support the view that traumatic experiences can have a causal effect on psychosis risk, it is important to understand the extent of gene-environment correlation in this relationship as: i) this will lead to more accurate estimates of any causal effect of trauma on psychosis, and ii) this can support the argument to provide support to individuals at high genetic risk to minimise the occurrence of traumatic events. To examine this possibility, we investigated genetic correlations between psychosis and trauma by testing the association between polygenic scores (PGS) for schizophrenia and childhood trauma exposure across two international longitudinal cohorts.

## Methods

### Avon Longitudinal Study of Parents and Children (ALSPAC)

ALSPAC is a longitudinal pregnancy cohort which aimed to recruit all pregnant women in the former county of Avon with an expected due date between April 1991 and December 1992. Detailed information has continued to be collected on mothers, partners and children in the cohort, this process has been described in detail elsewhere (Boyd *et al*., 2013; Fraser *et al*., 2013; Northstone *et al*., 2019). Ethics approval for the study was obtained from the ALSPAC Ethics and Law Committee and the Local Research Ethics Committees. Informed consent for the use of data collected via questionnaires and clinics was obtained from participants following the recommendations of the ALSPAC Ethics and Law Committee at the time. Please note that the study website contains details of all the data that is available through a fully searchable data dictionary and variable search tool.

### Measures of childhood trauma

The ALSPAC measures of childhood trauma were described in detail elsewhere (Croft *et al*., 2019). In brief, trauma exposure was collected prospectively from 0-17 years and supplemented by retrospective data collected at age 22 years. Information was collected on several categories of trauma exposure and questions were carefully selected to reflect experiences that would likely be highly upsetting to anyone encountering them. In ALSPAC, childhood trauma was derived using parent and child responses to a range of questionnaires collected across childhood and adolescence. In early childhood (before age 5 years), only parent-reported data were available, while in adolescence (between ages 11 and 17 years) measures of trauma were mainly child-reported. When both child- and parent-report data were available, these were combined to derive the exposure to trauma measure.

A composite measure of ‘any trauma’ was derived spanning the whole of childhood and adolescence (0-17 years) as well as trauma at several intervals (0-4.9 years, 5-10.9 years, and 11-17 years; Figure 1). Trauma exposures were also separated into distinct domains, namely bullying, domestic violence, sexual abuse, emotional neglect, emotional cruelty, and physical cruelty.

**Figure 1.**
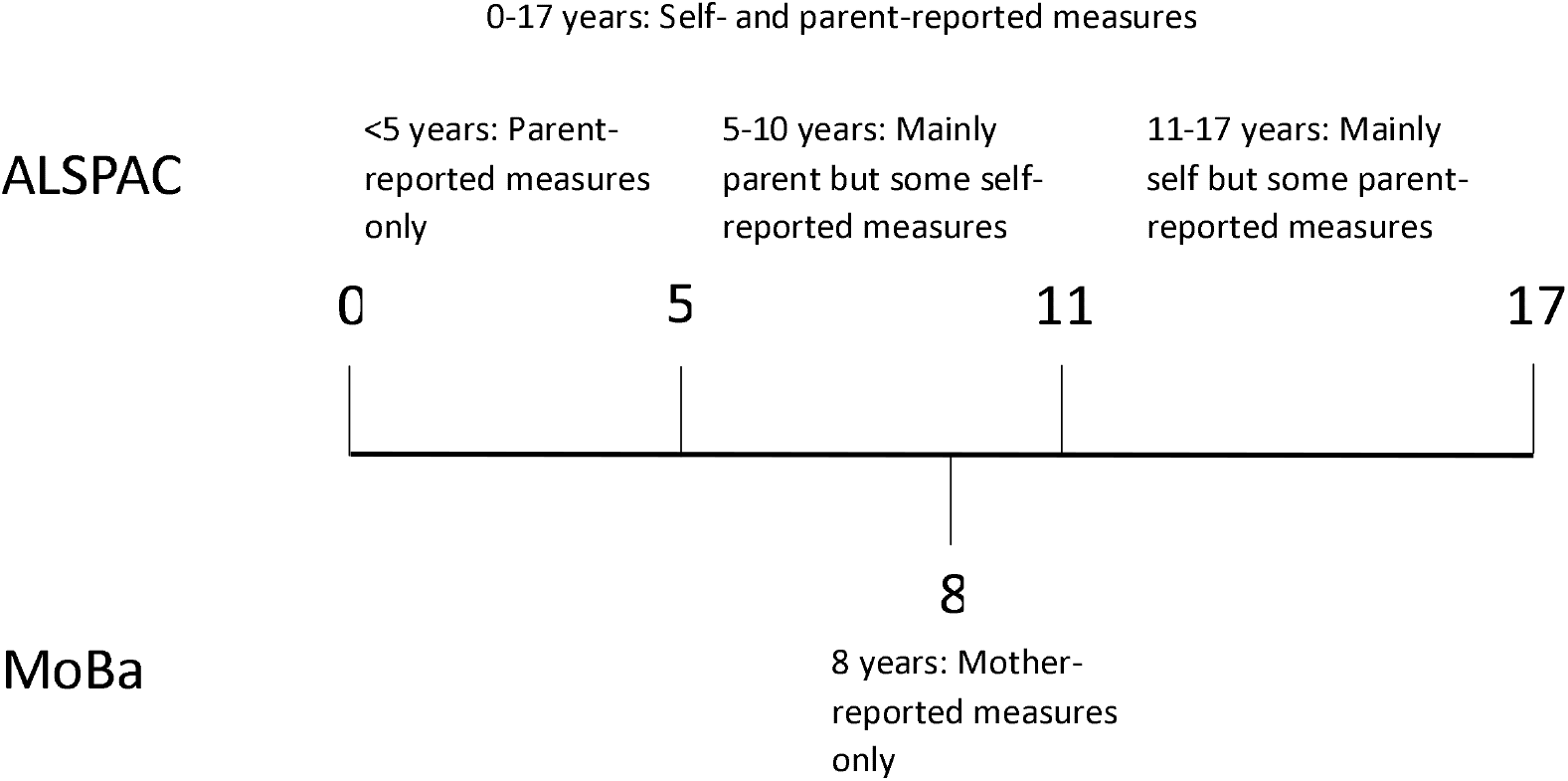
Timeline of data collection for the trauma measures in the Avon Longitudinal Study of Parents and Children (ALSPAC) and the Norwegian Mother, Father and Child Cohort Study (MoBa)

### Polygenic scores (PGS) for schizophrenia liability

Genotyped data were available on 7,977 children and 8,196 mothers in the ALSPAC study. Data were also available on a subset of fathers within the cohort (n=1,481). Details of genotyping and quality control measures are available in Supplementary Materials.

PGS for schizophrenia liability were derived for mothers, fathers, and children with genetic data available and were standardised prior to analyses. Schizophrenia PGS were derived as described in Jones et al. (2018). These scores were based on publicly available summary statistics published by the Psychiatric Genomics Consortium (PGC) (Schizophrenia Working Group of the Psychiatric Genomics Consortium, 2014). Overlapping SNPs in the GWAS summary statistics and the ALSPAC imputed genetic data were identified and LD clumping was used to identify independent genetic variants, with priority given to those with a stronger association in the PGC discovery GWAS. Weighted PGS were calculated using the effect estimates from the PGC GWAS, with SNPs being included in the PGS depending on the strength of the association in the original PGC GWAS. Thirteen schizophrenia association p-value thresholds were used ranging from p<0.5 to p<5×10^−8^ (see **Table S1** for a complete list), with a greater number of SNPs included as the p-value thresholds became less conservative.

### Statistical analyses

#### Multiple imputation

To maximise sample size and reduce selection bias due to attrition, multiple imputation was used to impute missing values in the trauma data. The imputation model included a broad range of variables related to trauma or variables known to be associated with sample missingness, in addition to the schizophrenia PGS and all variables included in any of the analysis models. A full description of the imputation strategy can be found in the Supplementary Materials.

#### Analysis

All analyses were performed in Stata 15 (StataCorp, 2017). Unadjusted logistic regression was used to investigate the strength of association between the schizophrenia PGS and experiencing childhood trauma. For our main analyses, we focused on PGS at a threshold of p_≤_0.05 (PGS_0.05_) as this threshold explains the most variability in genetic liability for schizophrenia in other samples (Schizophrenia Working Group of the Psychiatric Genomics Consortium, 2014).

Four main outcomes were examined; these were trauma measured across the whole of childhood and adolescence (age 0-17 years) and then at specific intervals during this period (0-4.9 years, 5-10.9 years, 11-17 years). The associations between schizophrenia PGS_0.05_ and specific trauma subtypes were also investigated. Each analysis was repeated using PGS_0.05_ derived for the mothers, fathers and the children.

#### Sensitivity analyses

We performed three sensitivity analyses. First, we examined the association between all PGS thresholds from p<0.5 to p<5×10^−8^ and exposure to trauma measured between 0-4.9 years of age. By restricting our analyses to this younger age group, it was expected that attrition would be minimal, and effects observed within this subset would be less prone to selection bias. Analyses were also performed using the complete-case data, to check consistency of results with the imputed dataset.

Second, we performed unadjusted logistic regression of child PGS_0.05_ on trauma exposure, restricting the analysis to only those children with data available on mother and father genotype. We then re-ran this model, adjusting for mother and father genotype to estimate the association between child PRS and trauma, independent of parental genotype.

Finally, we examined whether the association with trauma exposure was specific to genetic liability for schizophrenia or if it extended to psychiatric traits more generally. We created maternal and child PGS_0.05_ for several additional psychiatric phenotypes, described below, and conducted unadjusted logistic regressions to investigate the association between these scores with exposure to ‘any trauma’ at age 0-4.9 years. These additional phenotypes were attention-deficit hyperactivity disorder (ADHD) (Demontis *et al*., 2019), autism spectrum disorder (ASD) (Grove *et al*., 2019), bipolar disorder (Stahl *et al*., 2018), major depressive disorder (MDD) (Wray *et al*., 2018), neuroticism (Luciano *et al*., 2018), an updated schizophrenia risk score (Pardiñas *et al*., 2018), and a joint cross-disorder phenotype consisting of five major psychiatric disorders (Cross-Disorder Group of the Psychiatric Genomics Consortium *et al*., 2013).

### Replication in MoBa

We attempted to replicate the PGS-trauma association in an independent sample, the Norwegian Mother, Father and Child Cohort Study (MoBa). MoBa is a prospective population-based pregnancy cohort study conducted by the Norwegian Institute of Public Health (Magnus *et al*., 2016). Participants were recruited from all over Norway from 1999-2008. In 40.6 % of the pregnancies, women consented to participate. The cohort now includes 114,500 children, 95,200 mothers and 75,200 fathers. Blood samples were obtained from both parents during pregnancy and from mothers and children (umbilical cord) at birth. Genotyping is ongoing. The current study is based on version 12 of the quality-assured data files released for research in August 2018. The establishment and data collection in MoBa were previously based on a license from the Norwegian Data protection agency and approval from The Regional Committees for Medical and Health Research Ethics (REC); it is now based on regulations related to the Norwegian Health Registry Act. The current analyses were approved by REC (reference number 2016/1702).

Details of genotyping and quality control measures in MoBa are available in Supplementary Materials. PGS_0.05_ for schizophrenia liability were calculated for mothers, fathers and children with genetic data available at the time of analysis (15,208 children, 14,804 mothers and 15,198 fathers). Childhood trauma was measured in MoBa at age 8 years using maternal responses to questions selected to reflect experiences that would be highly upsetting to anyone encountering them (Figure 1). There were genetic data and trauma data available on 7,244 children. Bullying was defined as being subjected to beating, kicking or other violence by other children in the past year (n=1,665, 23.0%), emotional neglect as not usually letting the child know when he/she is doing a good job at something (n=124, 1.7%), and physical abuse as being subjected to beating, kicking or other violence by adults in the past year (n=49, 0.7%). In addition, the three questions were used to derive a composite measure of ‘any trauma’ (n=1,765, 24.3%). Details of the questions and prevalence in the full MoBa cohort are available in supplementary materials (Table S2).

Within MoBa, the association between PGS_0.05_ and trauma exposure was assessed in complete-cases using logistic regression adjusting for chip, batch and 10 principle components.

## Results

### Sample description

After imputation, 13,595 participants had data on childhood trauma within ALSPAC. By 17 years, 72.8% of the sample were reported to have experienced any trauma. Levels of trauma were lower in those with genetic data (n=9,946), with 70.7% of participants experiencing trauma compared to 78.5% of those without genetic data (n=3,649) (**Table 1**).

**Table 1.** Proportion with reported trauma across each age range in ALSPAC participants with and without genetic data

| Any childhood trauma | Whole sample<br>(N=13595) | With genetic data<br>(n=9946) | Without genetic data<br>(n=3649) | P-value |
| --- | --- | --- | --- | --- |
| 0-5 years | 29.2 | 27.2 | 34.5 | <0.001 |
| 6-11 years | 50.1 | 47.9 | 56.2 | <0.001 |
| 11-17 years | 47.9 | 46.1 | 52.7 | <0.001 |
| 0-17 years | 72.8 | 70.7 | 78.5 | <0.001 |

In MoBa, 42,236 participants had information on trauma measured at 8 years; of these individuals, 25.3% reported experiencing any trauma at this age, with ‘bullying’ being the most frequently endorsed exposure (Table S2).

### Association between schizophrenia PGS and experiencing trauma across childhood and adolescence

Within ALSPAC and across PGS scores derived based on child and mother data, we found evidence of an association between the schizophrenia PGS_0.05_ and increased exposure to childhood trauma at ages 0-5 years, 6-11 years, and 11-17 years, as well as across childhood and adolescence (0-17 years: OR_Child_=1.14, 95% CI: 1.08, 1.20, p=8.4×10^−6^; OR_Mother_=1.13, 95% CI: 1.06,1.20, p=8.5×10^−5^; **Table 2**). When using the smaller sample of fathers that had schizophrenia PGS_0.05_ available, the effect estimates were compatible with those observed using the mother and child PGS_0.05_, although there was no strong evidence of an association (0-17 years: OR_Father_=1.04, 95% CI: 0.92, 1.17, p=0.549; **Table 2**).

**Table 2.** Unadjusted association between schizophrenia PGS_0.05_ and exposure to any trauma in ALSPAC.

|  | Child PGS<br>(N=7,426) |  |  | Mother PGS<br>(N=7,380) |  |  | Father PGS<br>(N=1,215) |  |  |
| --- | --- | --- | --- | --- | --- | --- | --- | --- | --- |
| Age range | OR | 95% CI | P-value | OR | 95% CI | P-value | OR | 95% CI | P-value |
| 0-4.9 years | 1.15 | 1.08, 1.22 | $6.4 \times 10^{-6}$ | 1.18 | 1.12, 1.25 | $1.6 \times 10^{-8}$ | 1.12 | 0.97, 1.30 | 0.118 |
| 5-10.9 years | 1.07 | 1.01, 1.13 | 0.018 | 1.10 | 1.03, 1.16 | 0.002 | 1.06 | 0.95, 1.20 | 0.291 |
| 11-17 years | 1.13 | 1.07, 1.20 | $4.4 \times 10^{-5}$ | 1.15 | 1.08, 1.22 | $1.3 \times 10^{-5}$ | 0.99 | 0.87, 1.11 | 0.808 |
| 0-17 years | 1.14 | 1.08, 1.20 | $8.4 \times 10^{-6}$ | 1.13 | 1.06, 1.20 | $8.5 \times 10^{-5}$ | 1.04 | 0.92, 1.17 | 0.549 |

OR: odds ratio; PGS: polygenic score; CI: confidence interval

### Subtypes of trauma

When investigating the association between schizophrenia PGS_0.05_ and different types of trauma within ALSPAC, we found both maternal and child PGS_0.05_ were associated with the majority of trauma subtypes (**Table 3**). However, we found no robust evidence of an association between either maternal or child PGS_0.05_ and bullying. We also observed a similar pattern of effects when using paternal PGS_0.05_ (Table 3).

**Table 3.** Unadjusted association between schizophrenia PGS_0.05_ and subtypes of trauma across childhood and adolescence (age 0-17 years) in ALSPAC

|  | Child PGS<br>(N=7,426) |  |  | Mother PGS<br>(N=7,380) |  |  | Father PGS<br>(N=1,215) |  |  |
| --- | --- | --- | --- | --- | --- | --- | --- | --- | --- |
| Type of trauma | OR | 95% CI | P-value | OR | 95% CI | P-value | OR | 95% CI | P-value |
| Bullying | 1.03 | 0.97,1.09 | 0.342 | 1.01 | 0.96,1.07 | 0.620 | 1.03 | 0.92,1.17 | 0.587 |
| Domestic violence | 1.07 | 1.01,1.13 | 0.028 | 1.16 | 1.09,1.23 | 3.4x10<sup>-6</sup> | 1.07 | 0.92,1.25 | 0.385 |
| Sexual abuse | 1.15 | 1.03,1.29 | 0.012 | 1.12 | 1.00,1.25 | 0.043 | 1.11 | 0.89,1.38 | 0.343 |
| Emotional neglect | 1.06 | 0.96,1.17 | 0.235 | 1.11 | 1.01,1.22 | 0.033 | 0.84 | 0.68,1.04 | 0.115 |
| Emotional cruelty | 1.16 | 1.09,1.24 | 3.49x10<sup>-6</sup> | 1.15 | 1.08,1.24 | 6.7x10<sup>-5</sup> | 1.05 | 0.90,1.22 | 0.534 |
| Physical cruelty | 1.12 | 1.05,1.20 | 7.9x10<sup>-4</sup> | 1.16 | 1.08,1.24 | 1.7x10<sup>-5</sup> | 1.08 | 0.93,1.25 | 0.298 |

OR: odds ratio; PGS: polygenic score; CI: confidence interval

### Sensitivity analyses

When using both parental and child PGS generated using SNPs associated with schizophrenia at a range of p-value thresholds, we observed a positive association between increased PGS and experiencing trauma at all p-value thresholds (**Table S1**).

When looking at the association between PGS_0.05_ and trauma across the lifecourse using complete-case data, effect estimates were largest at the youngest ages where attrition was minimal (**Table S3**), and these were in a consistent direction, although attenuated, compared to the effects estimated when using the imputed data.

In the restricted sample of individuals with data on both mother and father genotype, we found strong evidence of an association between increased child PGS_0.05_ and trauma, with the observed effect sizes larger than in the analyses containing the full child sample (**Table 4**). After adjusting for both maternal and paternal genotype, the strength of evidence of association was reduced, although effect estimates were consistent with those estimated in the full child sample (**Table 4**).

**Table 4:** Association between child PGS_0.05_ and trauma at each time point – analysisrestricted to subset with maternal and paternal PGS

| Age range | Child PGS unadjusted<br>(N=1,526) |  |  | Child PGS adjusted*<br>(N=1,526) |  |  |
| --- | --- | --- | --- | --- | --- | --- |
|  | OR | 95% CI | P-value | OR | 95% CI | P-value |
| 0-5 years | 1.21 | 1.06, 1.39 | 0.005 | 1.09 | 0.92, 1.30 | 0.307 |
| 6-11 years | 1.10 | 0.98, 1.22 | 0.094 | 1.05 | 0.92, 1.21 | 0.474 |
| 11-17 years | 1.15 | 1.03, 1.29 | 0.015 | 1.14 | 0.99, 1.31 | 0.069 |
| 0-17 years | 1.15 | 1.03, 1.29 | 0.014 | 1.13 | 0.98, 1.30 | 0.096 |

\* Adjusted for maternal and paternal PGS<sub>0.05</sub>
OR: odds ratio; PGS: polygenic score; CI: confidence interval

We also examined the association of alternative psychiatric PGS_0.05_ with childhood trauma. We found strong evidence of an association between the MDD, neuroticism, cross-disorder phenotype and the updated schizophrenia PGS and childhood trauma, with similar effect sizes found for most of these associations (**Table S4**). For both cross-disorder and MDD, these associations were stronger when using the child PGS_0.05_ than the maternal PGS_0.05_. There was also some evidence of an association between childhood trauma and ADHD and bipolar disorder PGS_0.05_, but this was weaker. Evidence for these findings was consistent when using either maternal or child PGS_0.05_.

### Replication in an independent sample

Within MoBa, there was some evidence of association between the schizophrenia PGS_0.05_ and trauma exposures at age 8 years (**Table 5**). The effect estimates for any trauma were largely consistent with those estimated in ALSPAC at a similar age (6-11 years; **Table 2**).

**Table 5:** Association between PGS_0.05_ and exposure to trauma in the Norwegian Mother, Father and Child Cohort Study (MoBa)

|  | Child PGS<br>(N=7,244) |  |  | Mother PGS<br>(N=7,009) |  |  | Father PGS<br>(N=7,153) |  |  |
| --- | --- | --- | --- | --- | --- | --- | --- | --- | --- |
| Type of trauma | OR | 95% CI | P-value | OR | 95% CI | P-value | OR | 95% CI | P-value |
| Bullying | 1.09 | 1.03, 1.15 | 0.003 | 1.09 | 1.03, 1.15 | 0.003 | 1.07 | 1.01, 1.13 | 0.019 |
| Emotional neglect | 0.87 | 0.73, 1.04 | 0.122 | 1.12 | 0.93, 1.34 | 0.221 | 0.84 | 0.70, 1.00 | 0.046 |
| Physical abuse | 1.33 | 1.00, 1.76 | 0.047 | 0.96 | 0.72, 1.28 | 0.785 | 1.61 | 1.21, 2.13 | 0.001 |
| Any trauma | 1.08 | 1.02, 1.14 | 0.005 | 1.10 | 1.04, 1.16 | 0.001 | 1.06 | 1.00, 1.11 | 0.052 |

OR: odds ratio; PGS: polygenic score; CI: confidence interval

## Discussion

The main finding of this study is that PGS for schizophrenia liability were associated with increased exposure to childhood trauma within two independent and international birth cohorts. This finding was consistent when using both parental and child PGS, suggesting that it is unlikely to be solely driven by direct genetic effect from either parents or children. We also found that genetic liability to a range of additional psychiatric traits were associated with a greater trauma exposure. This highlights that individuals at higher genetic risk for poor mental health may benefit from supports that decrease their risk of trauma. It may also suggest that the original GWASs of these psychiatric traits are not only identifying variants associated with the trait of interest, but are also reflecting the effects of risk factors for these traits.

As GWAS studies become larger and thus statistically more powerful, they can identify variants of increasingly small effect that are robustly associated with the disorder being studied. For example, prior to 2014, GWAS had identified around 30 loci associated with schizophrenia (Schizophrenia Working Group of the Psychiatric Genomics Consortium, 2014). With the publication of a large meta-analysis performed by the Schizophrenia Working Group of the PGC (2014) this increased to 108 loci in 2014 and to 145 loci in 2018 (Pardiñas *et al*., 2018). However, with these gains in power comes the potential to detect small effects that are not direct genetic effects, but rather may reflect the effects of modifiable risk factors for the disorder under investigation (Gage *et al*., 2016). An example of this is the 2008 paper by Amos and colleagues who performed a GWAS of lung cancer, and in the process identified a variant located in the nicotinic receptor gene cluster *CHRNA5-A3-B4* (Amos *et al*., 2008). This locus is known to be robustly associated with smoking quantity, a well-known risk factor for lung cancer (Tobacco and Genetics Consortium, 2010; Ware, van den Bree and Munafò, 2011). It therefore seems possible that if exposure to childhood trauma is a risk factor for schizophrenia, then variants associated with trauma exposure could be picked up in the original schizophrenia GWAS. In addition to acting as an instrument for schizophrenia, this PGS could therefore be acting as a proxy for factors that could contribute to trauma exposure, and the association we observe between the PGS for schizophrenia liability and childhood trauma could be as a result of this. If sufficient data and robust instruments were to become available, approaches such as Mendelian randomisation for mediation could be used to contribute to these questions.

In addition to investigating the association with a single composite measure of trauma, we also examined several subdomains of trauma in an attempt to disentangle the relationship further. These subdomains were: bullying, domestic violence, sexual abuse, emotional neglect, emotional cruelty, and physical cruelty. When using both maternal and child PGS, we observed strong evidence of an association between increased schizophrenia PGS and all domains of childhood trauma across the life course, with the exception of bullying. Paternal PGS again generally showed a consistent direction of effect but, due to the smaller sample size, analyses were underpowered to detect effects. With the exception of bullying, each of our trauma subdomains are likely to occur within the home environment. Bullying, which often takes place at school and in the neighbourhood, showed little evidence of an effect in the ALSPAC cohort. Within ALSPAC, the bullying domain predominantly captured peer bullying, with only a single item on sibling bullying. In contrast, we did observe some association within MoBa, however, the bullying measure in MoBa includes exposure to violence from siblings. This suggests that there may be something specific to the home environment that is influencing this PRS-trauma exposure association, indicative of gene-environment correlation. If evocative gene-environment correlation was at play, we may expect to observe an association with bullying in addition to the other types of trauma. Equally, if active gene-environment correlation was occurring, we may expect to see a stronger association with the child PGS compared to the parental PGS. Given that we do not see either, and instead find that the association appears to be specific to more likely home-based exposures, it is possible that some form of passive gene-environment correlation is occurring, and that trauma could be a marker of genetic liability for later psychosis as well as a causal risk factor for psychosis. If the association that we observe in the home environment is due to parental traits, then establishing which of these traits play a role in shaping the greater ‘traumagenic’ environment could enable us to target modifiable factors in order to reduce their occurrence and impact. Although sufficient data is not currently available, analyses investigating between-sibling effects could go some way to disentangling these associations.

After adjusting analyses for parental PGS to investigate the effect of child genotype independently of parental genetic effects, we found that at younger ages the child genotype effect estimates attenuated substantially, while during adolescence effect estimates remained consistent. Since trauma is predominantly parent-reported at younger ages with an increasing number of self-reported measures in adolescence, one explanation is that this could be capturing reporting bias. An alternative argument is that child genotype becomes more influential for later trauma as children age, hence a transition occurs from a passive gene-environment correlation to a more active gene-environment correlation model. Previous findings suggest that those exposed to trauma in early life are at greater risk of revictimization in adolescence, which may also contribute to this observation (Fisher *et al*., 2015). If true, prevention measures should focus on parents and supporting a healthy homelife earlier on in childhood and targeting the children themselves as they approach adolescence. Developing prevention measures to reduce vulnerability to trauma among individuals with a higher genetic liability to poor mental health if timed right could help to reduce subsequent onset of mental illness among these high-risk individuals.

We also investigated the association between childhood trauma with child and maternal PGS for a number of alternative psychiatric phenotypes, using the latest available data (Pardiñas *et al*., 2018). We observed an association between several of these scores and childhood trauma; however, the strongest effects were with PGSs for schizophrenia and neuroticism. Results from the updated schizophrenia GWAS were comparable with the main analysis using PGS from the 2014 GWAS. There was strong evidence when looking at child PGS for MDD and a cross-disorder phenotype, but these effects were smaller when using maternal PGS. It seems likely that the association with childhood trauma is not necessarily specific to schizophrenia, but genetic predisposition to poor mental health in general. These findings are in line with recent work by Schoeler and colleagues (2019) and Leppert and colleagues (2019) who both found an association with genetic vulnerability to mental illness and exposure to bullying and other stressful life events.

The observed association between genetic liability for poor mental health and exposure to childhood trauma does not suggest that the majority of individuals with a mental health disorder will expose their children to traumatic events. The measure of trauma used in our study was very broad, meaning that exposure to it was common, even for individuals with a low genetic liability. The mechanisms through which the association between genetic liability and childhood trauma acts could be such that individuals with a higher genetic liability for poor mental health outcomes may end-up in more deprived neighbourhoods where exposure to trauma may be more common (Solmi *et al*., 2019), or that they engage in relationships where domestic violence is more likely to occur. These mechanisms would again suggest a form of passive gene-environment correlation, with the association arising between the genetic information passed from parent to child, and the environment in which the child is raised. If this is the case, then this suggests that those with higher genetic liability for mental illness may be most vulnerable to these effects and should be offered increased support to decrease their risk of trauma.

Another potential implication of our findings is that the observational association between childhood trauma and subsequent psychotic events may be, in part, due to genetic confounding, meaning that any causal effect of trauma on psychosis is somewhat weaker than the observed association between childhood trauma and subsequent psychosis would indicate. In a previous study using the ALSPAC cohort, we showed that the association between trauma and psychotic experiences was not attenuated by adjusting for either child or mother genetic risk for schizophrenia (Croft *et al*., 2019). Therefore, whilst the results from this present study indicate that exposure to trauma is higher where parents have a higher genetic risk for psychiatric disorders, this does little to challenge the theory that trauma has a causal effect on psychosis risk. Whilst measurement error in the PGS could have led to residual confounding in the study by Croft et al, and led to an overestimate of association, it seems unlikely that gene-environment correlation offers a plausible explanation for the trauma-psychosis relationship described in observational studies to date.

There are several limitations in our study that should be considered. First, there was substantial missing data in the trauma variable which could have resulted in selection bias. However, we used multiple imputation to impute our sample and maximise sample size, and the effect estimates in the complete cases at the youngest age bracket were comparable with those from the imputed dataset. In ALSPAC, attrition has been shown to be patterned by genetic risk factors (Taylor *et al*., 2018), therefore given the increased possibility of selection bias when using data at the older ages where attrition was greater, we also restricted this analysis to trauma measured up to age 5 years. We observed similar results at this age to the overall effect estimates, with a positive association between increased PGS and experiencing trauma. Second, the sub-set of fathers with genetic data was limited and so these father-based analyses were underpowered. We aimed to improve this by replicating in MoBa. Third, the trauma variables in the MoBa cohort were measured at age 8 years only and not as extensively as in the ALSPAC cohort. Therefore, we were unable to recreate completely comparable subdomains of trauma to disentangle the gene-environment correlation in this cohort. Fourth, in MoBa and the early childhood measures of trauma in ALSPAC were parent-reported only. It would be interesting to repeat these analyses comparing the parental PGS with parent-reported trauma exposure and child PGS with self-reported trauma exposure. However, in ALSPAC at the ages where self-reported measures were available (age 8 years onwards), the direction of effect remained consistent. Including analysis of parent’s own reported exposure to childhood trauma may also provide a more complete context for understanding the interplay between genetic risk and environmental factors that contribute to the risk of traumatic exposure during childhood.

## Conclusion

Analyses across two international birth cohorts indicate that genetic liability for schizophrenia, as well as other psychiatric phenotypes, is associated with childhood trauma. This could suggest that GWAS for these psychiatric traits are not only identifying variants associated with these traits, but may also reflect risk factors associated with them. We also found evidence to suggest that the association between genetic liability for schizophrenia and exposure to trauma has some specificity to the home environment and suggests that youth at higher genetic risk might require greater resources/support to ensure they grow-up in a healthy environment.

## Data Availability

The ALSPAC study website contains details of all the data that is available through a fully searchable data dictionary and variable search tool. These data are available on application to the ALSPAC exec.

## ALSPAC

We are extremely grateful to all the families who took part in this study, the midwives for their help in recruiting them, and the whole ALSPAC team, which includes interviewers, computer and laboratory technicians, clerical workers, research scientists, volunteers, managers, receptionists and nurses.

## MoBa

We thank the Principle Investigators of the major genotyping projects, Per Minor Magnus (NIPH) and Pål Njølstad (University of Bergen) for providing genotype data for the replication in MoBa. MoBa is supported by the Norwegian Ministry of Health and Care Services and the Ministry of Education and Research. We are grateful to all the participating families in Norway who take part in this on-going cohort study. We thank the Norwegian Institute of Public Health (NIPH) for generating genomic data in MoBa as part of the HARVEST collaboration, supported by the Research Council of Norway (NRC) (#229624). We also thank the NORMENT Centre for providing genotype data, funded by NRC (#223273), South East Norway Health Authority and KG Jebsen Stiftelsen. Further we thank the Center for Diabetes Research, the University of Bergen for providing genotype data and performing quality control and imputation of the data funded by the ERC AdG project SELECTionPREDISPOSED, Stiftelsen Kristian Gerhard Jebsen, Trond Mohn Foundation, NRC, the Novo Nordisk Foundation, the University of Bergen, and the Western Norway health Authorities (Helse Vest). We thank Laurie Hannigan (Lovisenberg Diaconal Hospital) for assisting with the QC and polygenic score analysis.

## References

Amos, C. I. et al. (2008) ‘Genome-wide association scan of tag SNPs identifies a susceptibility locus for lung cancer at 15q25.1’, Nature Genetics, 40(5), pp. 616–622. doi: 10.1038/ng.109.

Boyd, A. et al. (2013) ‘Cohort Profile: the ‘children of the 90s’--the index offspring of the Avon Longitudinal Study of Parents and Children’, Int J Epidemiol, 42(1), pp. 111–127. doi: 10.1093/ije/dys064.

Brumpton, B. et al. (2019) ‘Within-family studies for Mendelian randomization: avoiding dynastic, assortative mating, and population stratification biases’, bioRxiv. doi: 10.1101/602516.

Croft, J. et al. (2019) ‘Association of Trauma Type, Age of Exposure, and Frequency in Childhood and Adolescence With Psychotic Experiences in Early Adulthood’, JAMA Psychiatry. American Medical Association, 76(1), p. 79. doi: 10.1001/jamapsychiatry.2018.3155.

Cross-Disorder Group of the Psychiatric Genomics Consortium et al. (2013) ‘Genetic relationship between five psychiatric disorders estimated from genome-wide SNPs’, Nat Genet, 45(9), pp. 984–994. doi: 10.1038/ng.2711.

Cunningham, T., Hoy, K. and Shannon, C. (2016) ‘Does childhood bullying lead to the development of psychotic symptoms? A meta-analysis and review of prospective studies’, Psychosis. Routledge, 8(1), pp. 48–59. doi: 10.1080/17522439.2015.1053969.

van Dam, D. S. et al. (2012) ‘Childhood bullying and the association with psychosis in non-clinical and clinical samples: a review and meta-analysis’, Psychological Medicine. Cambridge University Press, 42(12), pp. 2463–2474. doi: 10.1017/S0033291712000360.

Davey Smith, G. and Ebrahim, S. (2005) ‘What can mendelian randomisation tell us about modifiable behavioural and environmental exposures?’, BMJ, 330(7499), pp. 1076–1079. doi: 10.1136/bmj.330.7499.1076.

Demontis, D. et al. (2019) ‘Discovery of the first genome-wide significant risk loci for attention deficit/hyperactivity disorder’, Nature Genetics. Nature Publishing Group, 51(1), pp. 63–75. doi: 10.1038/s41588-018-0269-7.

Fisher, H. L. et al. (2015) ‘Measuring adolescents’ exposure to victimization: The Environmental Risk (E-Risk) Longitudinal Twin Study’, Development and Psychopathology. doi: 10.1017/s0954579415000838.

Fraser, A. et al. (2013) ‘Cohort Profile: the Avon Longitudinal Study of Parents and Children: ALSPAC mothers cohort’, Int J Epidemiol, 42(1), pp. 97–110. doi: 10.1093/ije/dys066.

Gage, S. H. et al. (2016) ‘G = E: What GWAS Can Tell Us about the Environment’, PLOS Genetics, 12(2), p. e1005765. doi: 10.1371/journal.pgen.1005765.

Grove, J. et al. (2019) ‘Identification of common genetic risk variants for autism spectrum disorder’, Nature Genetics. Nature Publishing Group, 51(3), pp. 431–444. doi: 10.1038/s41588-019-0344-8.

Jaffee, S. R. and Price, T. S. (2008) ‘Genotype-environment correlations: implications for determining the relationship between environmental exposures and psychiatric illness.’, Psychiatry. NIH Public Access, 7(12), pp. 496–499. doi: 10.1016/j.mppsy.2008.10.002.

Jones, H. J. et al. (2018) ‘Investigating the genetic architecture of general and specific psychopathology in adolescence’, Translational Psychiatry. Nature Publishing Group, 8(1), p. 145. doi: 10.1038/s41398-018-0204-9.

Kong, A. et al. (2018) ‘The nature of nurture: Effects of parental genotypes’, Science. doi: 10.1126/science.aan6877.

Leppert, B. et al. (2019) ‘Association of Maternal Neurodevelopmental Risk Alleles With Early-Life Exposures’, JAMA Psychiatry. doi: 10.1001/jamapsychiatry.2019.0774.

Luciano, M. et al. (2018) ‘Association analysis in over 329,000 individuals identifies 116 independent variants influencing neuroticism’, Nature Genetics. Nature Publishing Group, 50(1), pp. 6–11. doi: 10.1038/s41588-017-0013-8.

Magnus P et al. (2016) ‘Cohort Profile Update: The Norwegian Mother and Child Cohort Study’, International Journal of Epidemiology, 45, pp. 382–8.

McGrath, J. J. et al. (2017) ‘Trauma and psychotic experiences: Transnational data from the World Mental Health survey’, British Journal of Psychiatry. doi: 10.1192/bjp.bp.117.205955.

Northstone, K. et al. (2019) ‘The Avon Longitudinal Study of Parents and Children (ALSPAC): an update on the enrolled sample of index children in 2019’, Wellcome Open Research, 4, p. 51. doi: 10.12688/wellcomeopenres.15132.1.

Pardiñas, A. F. et al. (2018) ‘Common schizophrenia alleles are enriched in mutation-intolerant genes and in regions under strong background selection’, Nature Genetics. Nature Publishing Group, 50(3), pp. 381–389. doi: 10.1038/s41588-018-0059-2.

Scarr, S. and McCartney, K. (1983) ‘How People Make Their Own Environments: A Theory of Genotype --> Environment Effects’, Child Development, 54(2), p. 424. doi: 10.2307/1129703.

Schizophrenia Working Group of the Psychiatric Genomics Consortium (2014) ‘Biological insights from 108 schizophrenia-associated genetic loci’, Nature, 511(7510), pp. 421–427. doi: 10.1038/nature13595.

Schoeler, T. et al. (2019) ‘Multi–Polygenic Score Approach to Identifying Individual Vulnerabilities Associated With the Risk of Exposure to Bullying’, JAMA Psychiatry. doi: 10.1001/jamapsychiatry.2019.0310.

Solmi, F. et al. (2019) ‘Neighborhood Characteristics at Birth and Positive and Negative Psychotic Symptoms in Adolescence: Findings From the ALSPAC Birth Cohort’, Schizophrenia Bulletin. doi: 10.1093/schbul/sbz049.

Stahl, E. A. et al. (2018) ‘Genome-wide association study identifies 30 Loci Associated with Bipolar Disorder’, bioRxiv. Cold Spring Harbor Laboratory, p. 173062. doi: 10.1101/173062.

StataCorp (2017) ‘Stata Statistical Software: Release 15’. College Station, TX: StataCorp LLC.

Taylor, A. E. et al. (2018) ‘Exploring the association of genetic factors with participation in the Avon Longitudinal Study of Parents and Children’, International Journal of Epidemiology. Narnia, 47(4), pp. 1207–1216. doi: 10.1093/ije/dyy060.

Tobacco and Genetics Consortium (2010) ‘Genome-wide meta-analyses identify multiple loci associated with smoking behavior’, Nature Genetics, 42(5), pp. 411–7. doi: 10.1038/ng.571.

Trotta, A., Murray, R. M. and Fisher, H. L. (2015) ‘The impact of childhood adversity on the persistence of psychotic symptoms: a systematic review and meta-analysis’, Psychological Medicine. Cambridge University Press, 45(12), pp. 2481–2498. doi: 10.1017/S0033291715000574.

Varese, F. et al. (2012) ‘Childhood Adversities Increase the Risk of Psychosis: A Meta-analysis of Patient-Control, Prospective- and Cross-sectional Cohort Studies’, Schizophrenia Bulletin. Oxford University Press, 38(4), pp. 661–671. doi: 10.1093/schbul/sbs050.

Ware, J. J., van den Bree, M. B. M. and Munafò, M. R. (2011) ‘Association of the CHRNA5-A3-B4 gene cluster with heaviness of smoking: a meta-analysis.’, Nicotine & tobacco researchlJ: official journal of the Society for Research on Nicotine and Tobacco. Oxford University Press, 13(12), pp. 1167–75. doi: 10.1093/ntr/ntr118.

Wray, N. R. et al. (2018) ‘Genome-wide association analyses identify 44 risk variants and refine the genetic architecture of major depression’, Nature Genetics, 50(5), pp. 668–681. doi: 10.1038/s41588-018-0090-3.

